# Fine-Grained Emotional Characterization of Dementia Caregivers in Online Support Communities Using Large Language Models

**DOI:** 10.64898/2026.07.26.26358983

**Authors:** Tushar Mungle, Audrey Angel Kwan, Yeon Mi Hwang, Malvika Pillai, Michelle Sahai, Madelena Ng, Rebecca Handler, Tina Hernandez-Boussard

**Author notes:** Correspondence to, Dr. Yeon Mi Hwang, PhD, Department of Medicine, Stanford University, 3180 Porter Drive, Palo Alto, CA, 94304, Dr. Tushar Mungle, PhD, Department of Medicine, Stanford University, 3180 Porter Drive, Palo Alto, CA, 94304. Equal contribution (Tushar Mungle, Audrey Angel Kwan, Yeon Mi Hwang).

## Abstract

**Background:** Dementia caregiving carries substantial emotional and psychological consequences, but most evidence comes from structured surveys and interviews that are resource-intensive and may incompletely capture spontaneous, contextual experience. Scalable methods to characterize caregiver experience from real-world narratives are lacking.

**Objective:** To characterize how emotional expression among dementia caregivers varies by caregiving relationship and context, using structured extraction of caregiver narratives from an online support community at scale.

**Methods:** We analyzed 7,198 publicly available posts from 3,350 authors across three forums of an online dementia caregiver community. A large language model (LLM) extracted caregiver role and relationship, caregiving objective, emotional valence (-1 to +1), and emotional themes from each post. Reproducibility of the extracted annotations was assessed through agreement between two independent LLMs, using Cohen’s κ for categorical fields and correlation for continuous valence.

**Results:** Caregiver narratives were predominantly negative (89.3% of posts; mean valence - 0.46), with interpretable structure. Parent caregivers (-0.51) and adult children caring for fathers (-0.52) and mothers (-0.50) expressed more negative valence than spouse caregivers (-0.47). Emotional valence varied most by post objective - most negative for immediate safety/crisis (-0.73), family conflict (-0.61), and end-of-life (-0.55) contexts, and net-positive only for resource sharing (+0.05). Even net-positive posts often retained concern alongside hope or gratitude rather than expressing uniform positivity. Concern, frustration, and sadness were the most prevalent emotional themes. Greater cumulative posting activity was associated with more positive expression. Inter-model agreement was high for relationship category (κ=0.80) and emotional valence (r=0.95).

**Conclusions:** Caregiver emotional expression is systematically patterned by caregiving relationship and by the objective of a post, with crisis and family conflict situations most negative and resource sharing the only net positive context, in ways that coarse sentiment or topic-modeling approaches have not shown. Emotional valence reflects expressed experience rather than clinical burden. Applied at scale, structured LLM extraction complements survey-based methods and could support distress screening and longitudinal monitoring of caregiver experience and inform the design of caregiver-support programs.

## Introduction

Family caregivers of persons living with Alzheimer’s disease and related dementias (ADRD) provide substantial support for activities of daily living, behavioral supervision, symptom management, healthcare coordination, and long-term decision-making responsibilities.^1^ Caregivers enable people with ADRD to remain in the community but often at the expense of substantial emotional and psychological consequences to their own well-being. This can include elevated stress, anxiety, depression, grief, social isolation, and reduced quality of life which intensify as people with ADRD experience more neuropsychiatric symptoms, cognitive decline, and increasing care demands.^2–4^ Although these consequences are well documented, most evidence derives from structured surveys, interviews, and cross-sectional assessments that capture predefined constructs at fixed time points.^4–7^ Such instruments are essential and well validated, but they are resource intensive to administer at scale and may incompletely capture the spontaneous, contextual, and often nuanced emotional experiences of caregiving as they naturally unfold. Self-reporting under structured conditions can also be shaped by recall and social-desirability effects. These constraints point to a need for complementary, scalable methods that can characterize caregiver experience organically.

Online caregiver communities offer such an opportunity. These communities represent a large and growing population of caregivers actively seeking connection with others navigating similar circumstances, a practical relevance for the design of caregiver-support programs and services. Caregivers use these forums to seek emotional support, exchange coping strategies, and describe caregiving challenges in their own words, generating a continuous stream of unfiltered narratives.^8,9^ As the participation is voluntary and frequently anonymous, these platforms capture spontaneous expression that may not surface under structured conditions and serve as a complement to traditional surveys.^10^ Prior work has demonstrated the feasibility of analyzing such data with natural language processing (NLP) to surface caregiver stressors, sentiment, and interaction patterns.^10–12^ However, these approaches have largely relied on topic modeling, lexicon-based sentiment, or coarse polarity categories (positive, neutral, negative). They reduce rich experience to broad labels and struggle to represent context, implicit distress, co-occurring (mixed) emotions, and the relational structure of who is caregiving and what they are seeking.^10–12^

What remains missing is a scalable way to convert these narratives into structured, multidimensional representations of caregiver experience that can be analyzed at population scale. Large language models (LLMs) make this feasible: through context-aware semantic interpretation, they can jointly extract latent constructs such as a caregiver’s role and relationship, the objective of a given account, and its emotional valence and themes from free text.^13,14^ Although emerging studies have applied LLMs to dementia care for caregiver support, information-needs assessment, and clinical knowledge extraction,^15–17^ LLMs have rarely been used to derive structured caregiver representations from real-world narratives at scale. Relationships are a particularly under-examined dimension: spouses, adult children, and other family caregivers differ in responsibilities, emotional proximity, and decision-making roles, yet prior narrative analyses seldom model relationship, caregiving objective, and emotional expression together.^18,19^

In this study, we ask how emotional expression among ADRD caregivers varies by caregiving relationship and by the purpose of a post, using structured extraction of a large corpus of narratives from an online caregiver community. We assess the reproducibility of the extracted annotations through agreement between independent LLMs and use the resulting structured data to characterize how emotional expression relates to caregiver relationship, caregiving objective, and community engagement. This approach is intended to complement established survey- and interview-based methods and to support downstream applications such as distress screening and longitudinal monitoring.

## Methods

### Data Source

Data were collected from AlzConnected (https://alzconnected.org), an online caregiver support community for individuals caring for persons living with Alzheimer’s disease and related dementias (ADRD). Publicly available discussion threads were extracted from three caregiver forums representing (a) general caregivers, (b) spouses/partners, and (c) caregivers of parents using an automated web-scraping pipeline. This process was completed with an asynchronous web scraping Python script, using aiohttp to make network requests and asyncio to schedule scraping tasks. The forums’ HTML was parsed with BeautifulSoup to dynamically construct URLs with page numbers to navigate between the forums’ pages and posts. Retry logic with delays was additionally implemented to handle cases where page and post links did not immediately load. BeautifulSoup and CSS selectors were then used to extract structured data from each post, including post titles, post content, author identifiers, posting dates, and discussion tags. Finally, after all pages were processed, the results were flattened into a single CSV dataset per forum.

### Data Processing

A multi-stage preprocessing pipeline was developed to improve dataset quality. Posts were first chronologically ranked and restricted to the most recent entries within each forum cohort (maximum 3,000 posts per forum) as of February 2, 2026, to capture recent caregiving experiences and reflect current care practices and discourse around dementia. Each post was then assigned a unique Original Post Identifier (OPID) to enable tracking across preprocessing, annotation, and integration stages.

Caregiver-specific abbreviations and ADRD terminology commonly used within the AlzConnected community were standardized prior to analysis. Frequently used shorthand expressions and caregiving terminology (e.g., AD, ADRD, PLWD, EOAD, MCI, LTC, SNF, POA) were identified from community-supported abbreviation resources available through the AlzConnected platform (https://alzconnected.org/discussion/64407/common-abbreviations) and expanded into standardized clinical terminology to improve semantic consistency.

To preserve participant privacy, a hybrid de-identification framework combining named entity recognition (NER) and rule-based masking was implemented. General entities including person names, organizations, dates, times, and geographic locations were identified using the spaCy en_core_web_sm model, whereas biomedical entities were detected using the en_ner_bc5cdr_md clinical NER model. Additional rule-based dictionaries and regular expressions were applied to remove dementia-related diagnoses and medication mentions. This step was undertaken as a privacy-protective safeguard prior to storage and analysis rather than a requirement of the data source, since the posts analyzed were publicly available and did not constitute protected health information. De-identified records were subsequently integrated across caregiver groups to create the unified analytical dataset.

### LLM-based Caregiver Information Extraction and Annotation

The central component of this study is a framework that transforms unstructured caregiver narratives into structured, multidimensional caregiver representations. Following preprocessing, de-identified posts were annotated using a multi-stage large language model (LLM) extraction pipeline operating on a secure infrastructure.^20^ Annotation used Claude-3.5-sonnet-20241022 (referred to as Claude in the remaining manuscript) with low-temperature, near-deterministic settings (temperature = 0.0, top-p = 0.25, maximum output = 500 tokens) to improve reproducibility.

Relationship specific prompting pipelines were developed for the general, spouse, and parent cohorts to reflect differences in caregiving roles and relationship definitions. Each prompt (Supplemental Methods M1) followed a structured format comprising (a) task instructions and annotation objectives, (b) a predefined JSON schema specifying permissible outputs and categories, and (c) contextual post information (title, content, author metadata, posting history, timestamps, and discussion tags). Model outputs were required to conform to the JSON schema, and both raw responses and parsed annotations were retained to preserve traceability between generated outputs and analytic variables.

The framework extracted the following dimensions for each post:

1. **Caregiver status:** non-caregiver, former caregiver, secondary caregiver, or primary caregiver (Prompts P1, P4, P5). Posts classified as non-caregiver or former caregiver were excluded from downstream analyses to retain only current caregiver narratives.
2. **Caregiver relationship:** relationship category (e.g., caring for spouse, adult childcaring for parent, other) and, where applicable, a more specific role (Prompts P1, P4, P5). Because the specific role (e.g., son vs. daughter) requires the post to state gender explicitly, this field is inherently lower yield and frequently resolves to “uncertain.”
3. **Caregiving objective:** a single primary objective selected from eleven predefined categories (Prompt P2), developed by the study team (authors - TM, AAK and YMH) through iterative review of forum content and consultation of prior caregiving-needs taxonomies to capture the primary reasons caregivers post to the forum (e.g., informational, emotional, or crisis-related needs).
4. **Emotional valence and themes:** a continuous emotional valence score from −1 (most negative) to +1 (most positive), three continuous sentiment probabilities (negative, neutral, positive) summing to 1, and up to three dominant emotional themes with supporting text (Prompt P3).

Dominant emotional themes were generated as free-text labels, prompted using Parrott’s hierarchical emotion taxonomy (2001) as a conceptual reference rather than a closed vocabulary; the resulting labels were therefore open-vocabulary emotion descriptors informed by, but not constrained to, Parrott’s categories. For each post, a binary sentiment classification was derived from the sign of the continuous emotional valence score (negative if < 0, positive if > 0).

### Reproducibility Assessment

To assess the reproducibility of the extracted annotations, agreement was computed between independently generated outputs from Claude and GPT-4o across the general, spouse, and parent cohorts, using identical prompts, contextual inputs, and inference settings to minimize procedural variability. Categorical fields (e.g., caregiver status, caregiving objective, relationship category and specific role) were evaluated using Cohen’s κ and exact percent agreement, interpreted using conventional thresholds (<=0.20 slight, 0.21-0.40 fair, 0.41-0.60 moderate, 0.61-0.80 substantial, >=0.80 almost perfect). The continuous emotional valence score was evaluated using Pearson and Spearman correlations and the mean absolute difference between models.

We assessed consistency through inter-model agreement using independent LLMs, following the view that reliability is a necessary first step toward validity in subjective domains where ground truth is often unattainable.

### Engagement (Frequent Contributor) Analysis

User engagement was operationalized as each author’s total cumulative post count on the platform obtained from author profile metadata. This measure is distinct from the number of an author’s posts contained in the analytic sample. Authors were stratified into quartiles by cumulative post count, and those in the upper quartile (>=157 posts) were classified as frequent contributors. This stratification was used to examine whether sustained, high-volume engagement is associated with distinctive patterns in caregiving objectives, relationship composition, and emotional expression. Because frequent-contributor status is an author-level attribute applied at the post level, post-level analyses involving this variable are subject to within-author clustering, which is addressed in the statistical analysis below.

### Statistical Analysis

Categorical variables are summarized as counts and percentages, and emotional valence as means with standard deviations and ranges. Differences in mean valence between two groups were tested using Welch’s t-test, and across three or more groups using one-way ANOVA; associations among categorical variables were tested using the χ^2^ test. Given the large sample size, effect sizes are reported alongside p-values and serve as the primary basis for interpretation: Hedges’ g (or Cohen’s d) for mean differences and Cramér’s V for χ^2^ associations; valence differences smaller than threshold, e.g., 0.05 valence units are treated as negligible regardless of statistical significance.

The association between cumulative posting activity and emotional valence was assessed at the author level, using one aggregate valence value per author, with Pearson and Spearman correlations. Binned summaries of valence by posting frequency are presented for visualization only and were not used for inference.

### Ethics Statement

This study was approved by the Stanford University Institutional Review Board under expedited review (eProtocol #83818). The study used publicly accessible discussion posts obtained from the AlzConnected caregiver forum, which does not require login or registration to access, and did not involve direct participant interaction or access to identifiable private information. All records underwent de-identification prior to analysis to further preserve participant privacy and confidentiality.

## Results

### Study Sample

A total of 9,627 caregiver posts were scraped from AlzConnected. The posts were sorted by date and only the most recent 3,000 posts per forum were kept. Posts classified as having caregiver statuses of “Not a caregiver” or “Former caregiver” were also excluded from downstream analyses. Thus, a total of 7,198 caregiver posts generated by 3,263 unique authors were analyzed between June 2022 and February 2026.

### Comparison of Traditional NLP and LLM-Based Emotional Assessment

The two approaches produced markedly different emotional distributions (Figure S1). Transformer-based sentiment scores were strongly bimodal, concentrating near the extreme-negative bound (sentiment ≤ −0.98) (Figure S1a) with a smaller cluster near neutral, a pattern consistent with the probability weighted scoring used to derive the continuous score. LLM-derived emotional valence for the same posts was more dispersed, extending across the full range from strongly negative to positive (Figure S1b). Both methods yielded a net negative mean. Because no human reference standard was available for this comparison, these results indicate that the two methods characterize emotional content differently, LLM assigning more graded, less polarized scores, rather than establishing that either is more accurate.

### Inter model Agreement

Agreement between Claude and GPT-4o annotations was assessed to evaluate reproducibility. Categorical fields ranged from fair to almost-perfect agreement: relationship category showed almost-perfect agreement (κ = 0.80; 83.8% exact agreement), caregiving objective substantial agreement (κ = 0.67; 73.4%), caregiver status moderate agreement (κ = 0.59; 76.0%), and relationship-specific role only fair agreement (κ = 0.33; 64.6%), consistent with the difficulty of inferring a caregiver’s gender-specific role from text. The continuous emotional valence score was highly reproducible across models (Pearson r = 0.95, Spearman ρ = 0.95, mean absolute difference = 0.11).

### Descriptive Characteristics

The study cohort comprised of 7,583 posts from 3,350 unique authors, drawn from the general (31.5%), spouse (34.8%), and parent (33.8%) forums (Table 1). Across all posts, the mean emotional valence was -0.46 (range -0.95 to +0.95). Most posts were classified as negative: 6,771 (89.3%) carried a negative valence and 807 (10.6%) a positive valence, with the remaining 5 posts having a neutral (zero) score (Table 1).

**Table 1:**
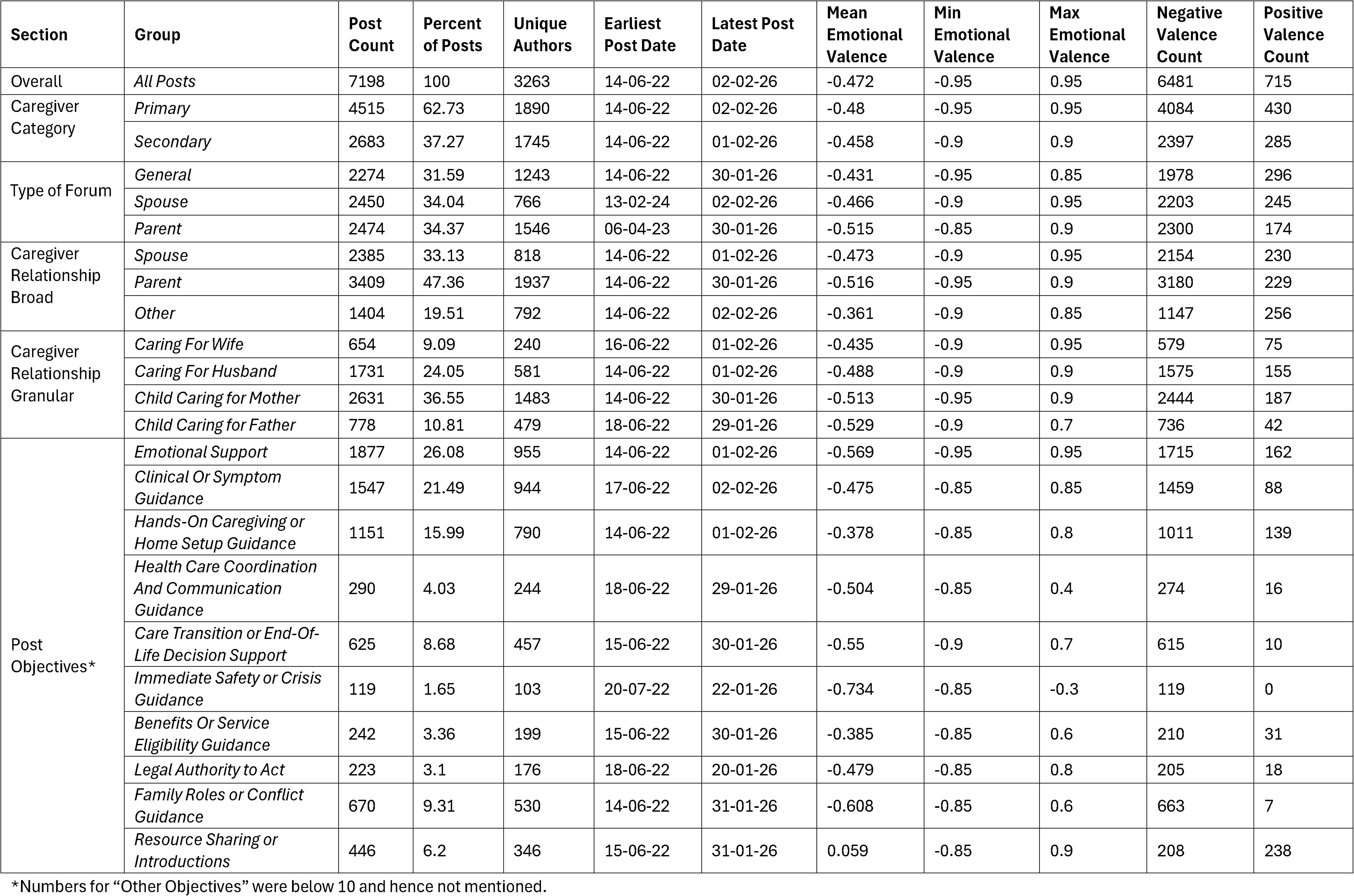
Characteristics of caregiver posts and emotional valence across caregiver groups, relationship categories, and caregiving objectives.

By caregiver category, primary caregivers contributed the largest share of posts (59.5%; mean valence -0.48), followed by secondary caregivers (35.4%; mean valence -0.46). By relationship, parent-caregiving was the most common category (46.8%) and showed the most negative mean valence (-0.51), followed by spouse caregiving (33.8%; -0.47); the heterogeneous “other” category - comprising grandparent, in-law, sibling, friend, and other-relative caregivers (19.5%), showed the least negative valence (-0.35) (Table 1). At the granular level, adult children caring for fathers showed the most negative mean valence (-0.52), followed by adult children caring for mothers (-0.50), caregivers of husbands (-0.48), and caregivers of wives (-0.43).

Across caregiving objectives, emotional-support requests were the most frequent (28.1%), followed by clinical or symptom guidance (20.5%) and hands-on caregiving or home setup guidance (15.4%) (Table 1). Mean valence varied substantially by objective: it was most negative for immediate safety or crisis guidance (-0.73; 121/121 posts negative) and family-roles or conflict guidance (-0.61), followed by care-transition or end-of-life decision support (-0.55) and emotional support (-0.54). Resource sharing or introductions was the only substantive objective category with a net-positive mean valence (+0.05; 262 of 502 posts, 52.2%, positive).

### Caregiving Objectives and Emotional Valence Across Caregiver Groups

Caregiving objectives differed substantially across caregiver categories and relationship structures (Figure 1). Among both primary and secondary caregivers, emotional support represented the dominant objective, accounting for 1,391 primary caregiver posts and 486 secondary caregiver posts, followed by clinical guidance (954 vs. 593 posts) and hands-on caregiving support (722 vs. 429 posts) respectively (Figure 1A). Primary caregivers consistently showed higher engagement across emotionally intensive domains including emotional support, end-of-life decision support, and symptom management.

**Figure 1.**
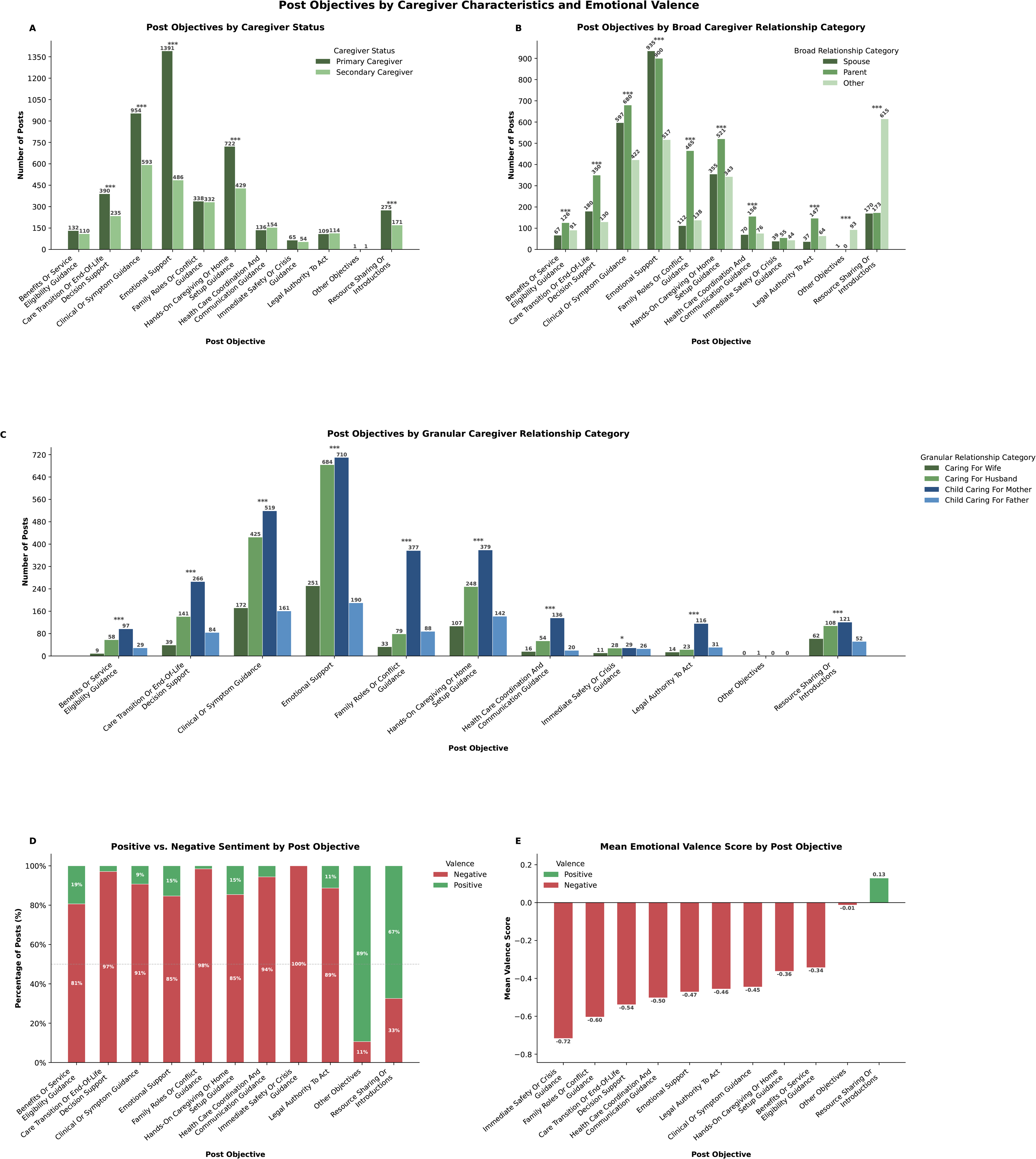
Distribution of post objectives and associated emotional expression across caregiver groups.(A) Distribution of primary caregiving objectives among primary and secondary caregivers. (B) Distribution of caregiving objectives across broad caregiver relationship categories (spouse, parent, and other caregivers). (C) Distribution of caregiving objectives across granular caregiver relationship categories (caring for wife, caring for husband, childcaring for mother, and childcaring for father). (D) Percentage of posts classified as positive or negative across caregiving objectives. (E) Mean emotional valence scores (range: -1 to +1) across caregiving objectives. Emotional valence was derived from LLM-based emotional profiling. Statistical significance was assessed using chi-square tests for categorical comparisons. p<0.05, *p<0.01, **p<0.001.

In relationship specific analyses (Figure 1B), parent-caregivers contributed the most posts on family-roles or conflict guidance (n = 465), healthcare coordination (n = 156), and legal planning (n = 147), whereas spouse caregivers were more strongly represented in emotional support (n = 935) and symptom-management requests (n = 597). Adult children caring for mothers were the largest granular subgroup and contributed the most emotionally oriented posts, including 710 emotional-support, 519 clinical-guidance, and 377 family-conflict posts (Figure 1C).

Emotional valence varied markedly by objective (Figure 1D, E). Immediate safety or crisis guidance had the most negative profile (mean valence -0.73; 100% negative posts), followed by family-roles or conflict guidance (-0.61; 98.5% negative) and care-transition or end-of-life decision support (-0.54; 97.1% negative). Resource-sharing was the only objective associated with predominantly positive expression (mean valence +0.05; 52.2% positive).

### Emotional Valence and Post Sentiment by Caregiver Characteristics

Primary caregivers demonstrated significantly greater emotional burden than secondary caregivers (Figure 2A), with lower mean emotional valence scores (-0.48 vs. -0.46, Welch p=0.011). However, sentiment distributions remained comparable between groups (Figure 2B), with approximately 90% negative posts observed in both primary (90.5%) and secondary caregivers (89.4%).

**Figure 2.**
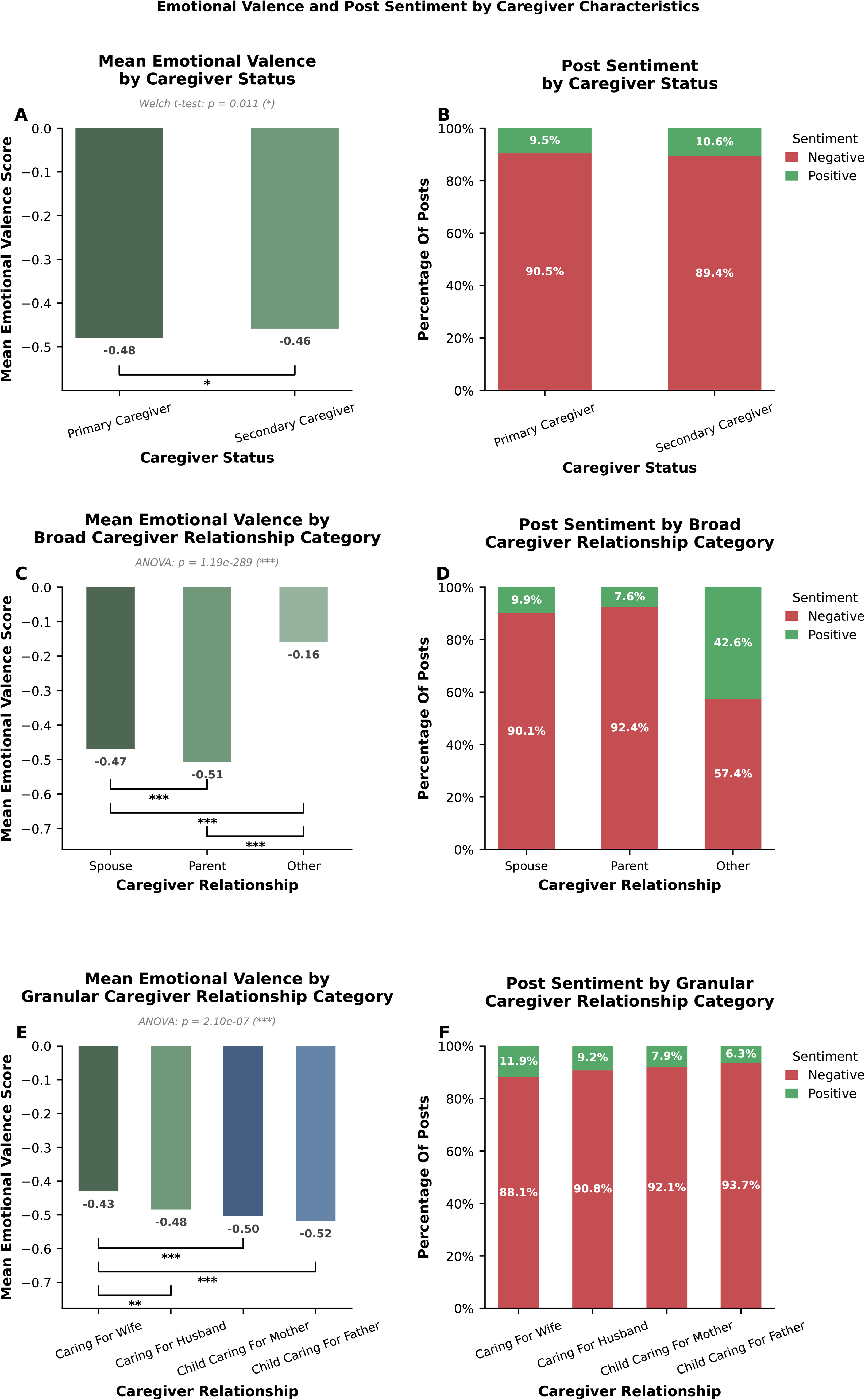
Emotional expression and sentiment distributions across caregiver categories and relationships. (A) Mean emotional valence scores among primary and secondary caregivers. (B) Distribution of positive and negative sentiment among primary and secondary caregivers. (C) Mean emotional valence scores across broad caregiver relationship categories. (D) Distribution of positive and negative sentiment across broad caregiver relationship categories. (E) Mean emotional valence scores across granular caregiver relationship categories. (F) Distribution of positive and negative sentiment across granular caregiver relationship categories. Emotional valence scores ranged from -1 (most negative) to +1 (most positive). Statistical comparisons were performed using Welch’s t-test (A) and one-way ANOVA (C, E). p<0.05, *p<0.01, **p<0.001.

Relationship level analyses showed substantial heterogeneity in emotional burden. Parent caregivers exhibited strong negative emotional valence (-0.51) followed by spouse caregivers (-0.47), whereas other caregiver relationships showed comparatively lower burden (-0.16) (Figure 2C). Parent caregivers additionally demonstrated the highest proportion of negative posts (92.4%) compared with spouses (90.1%) and other caregivers (57.4%) (Figure 2D). At the granular level, child caregivers supporting fathers showed the greatest emotional burden (Figure 2E) with mean valence scores of -0.52, followed by child caregivers supporting mothers (-0.50). Consistent trends were observed for sentiment distributions (Figure 2F), where child caregivers supporting fathers demonstrated the highest proportion of negative posts (93.7%), whereas caregivers supporting wives showed comparatively lower burden (88.1% negative).

### Emotional Theme Analysis Across Posts

Theme analysis identified concern (n=3,469) as the most prevalent emotional construct across caregiver narratives (Figure 3A), followed by frustration (n=2,506), sadness (n=2,039), uncertainty (n=1,522), and anxiety (n=1,495). Less frequent but clinically relevant themes included guilt and distress (<1000). Negative posts (Figures 3B and 3D) were dominated by concern, frustration, and sadness, whereas positive posts (Figure 3C and 3E) more frequently demonstrated hope, interest, gratitude, and concern.

**Figure 3.**
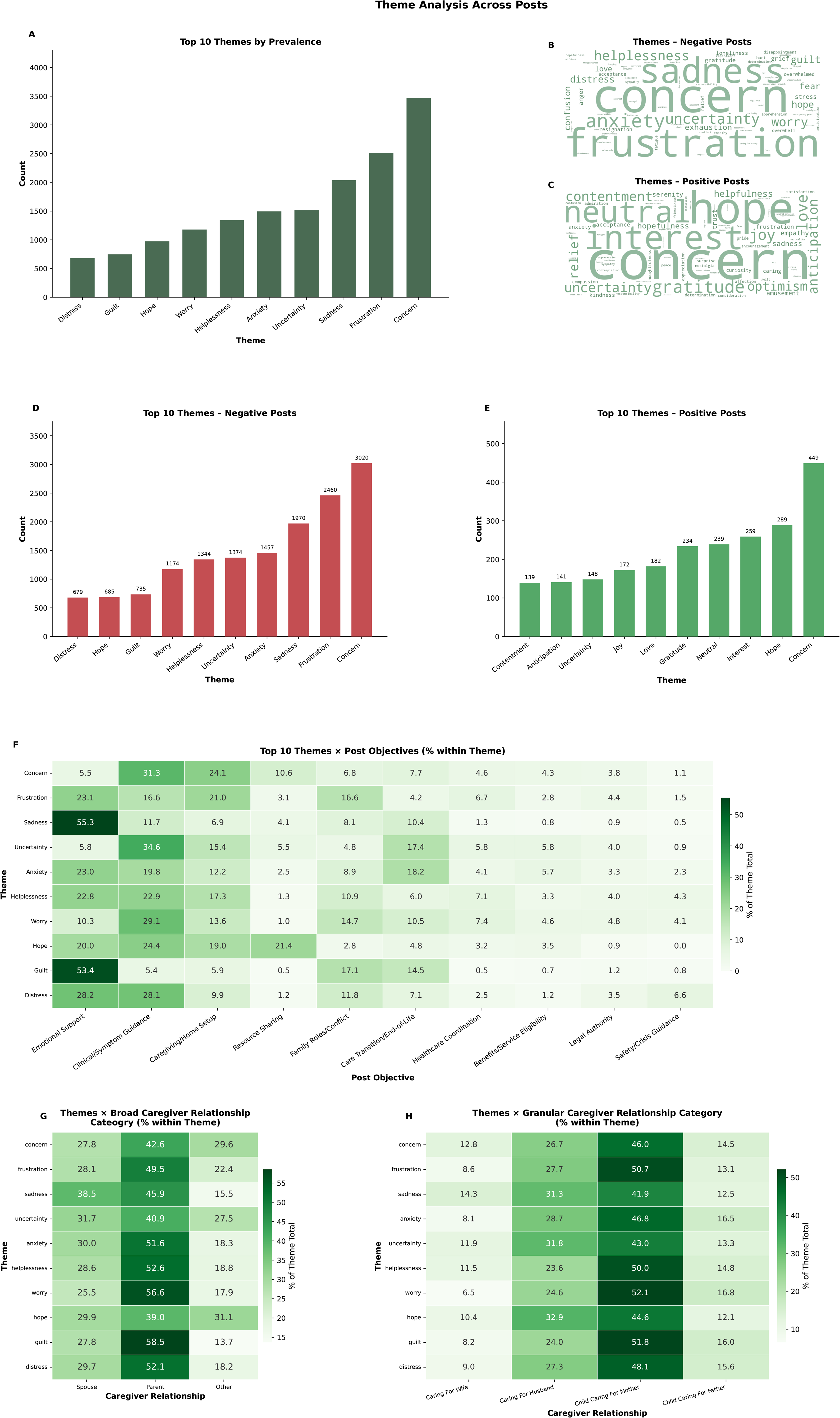
Distribution of emotional themes identified from caregiver narratives. (A) Ten most prevalent emotional themes across all caregiver posts. (B) Word cloud visualization of emotional themes identified in negative posts. (C) Word cloud visualization of emotional themes identified in positive posts. (D) Ten most prevalent emotional themes among negative posts. (E) Ten most prevalent emotional themes among positive posts. (F) Heatmap showing the distribution of emotional themes across caregiving objectives, expressed as the percentage contribution within each emotional theme. (G) Distribution of emotional themes across broad caregiver relationship categories. (H) Distribution of emotional themes across granular caregiver relationship categories. Emotional themes were derived using LLM-based annotation and mapped to Parrott’s hierarchical emotion framework. Values in heatmaps represent percentages within each emotional theme.

Emotional themes were strongly associated with caregiving objectives (Figure 3F; χ^2^=4957, p<0.001). Sadness and guilt clustered primarily within emotional support discussions, whereas concern, uncertainty and worry were concentrated in symptom management posts. Relationship analyses showed higher proportions of negative themes particularly for worry, helplessness, guilt, and distress as compared to spouse, among parent caregivers (Figure 3G). At granular level, adult children caring for mothers contributed the largest share of most theme categories, frequently accounting for 45-50% of theme’s total prevalence (Figure 3H).

### Posting Behavior and Contributor Characteristics

Posting frequency showed a strong positive relationship with emotional recovery (Figure 4A). Mean emotional valence progressively improved with increasing author activity, demonstrating a significant positive correlation between post count and emotional expression (*r*=0.82, *p*<0.001).

**Figure 4.**
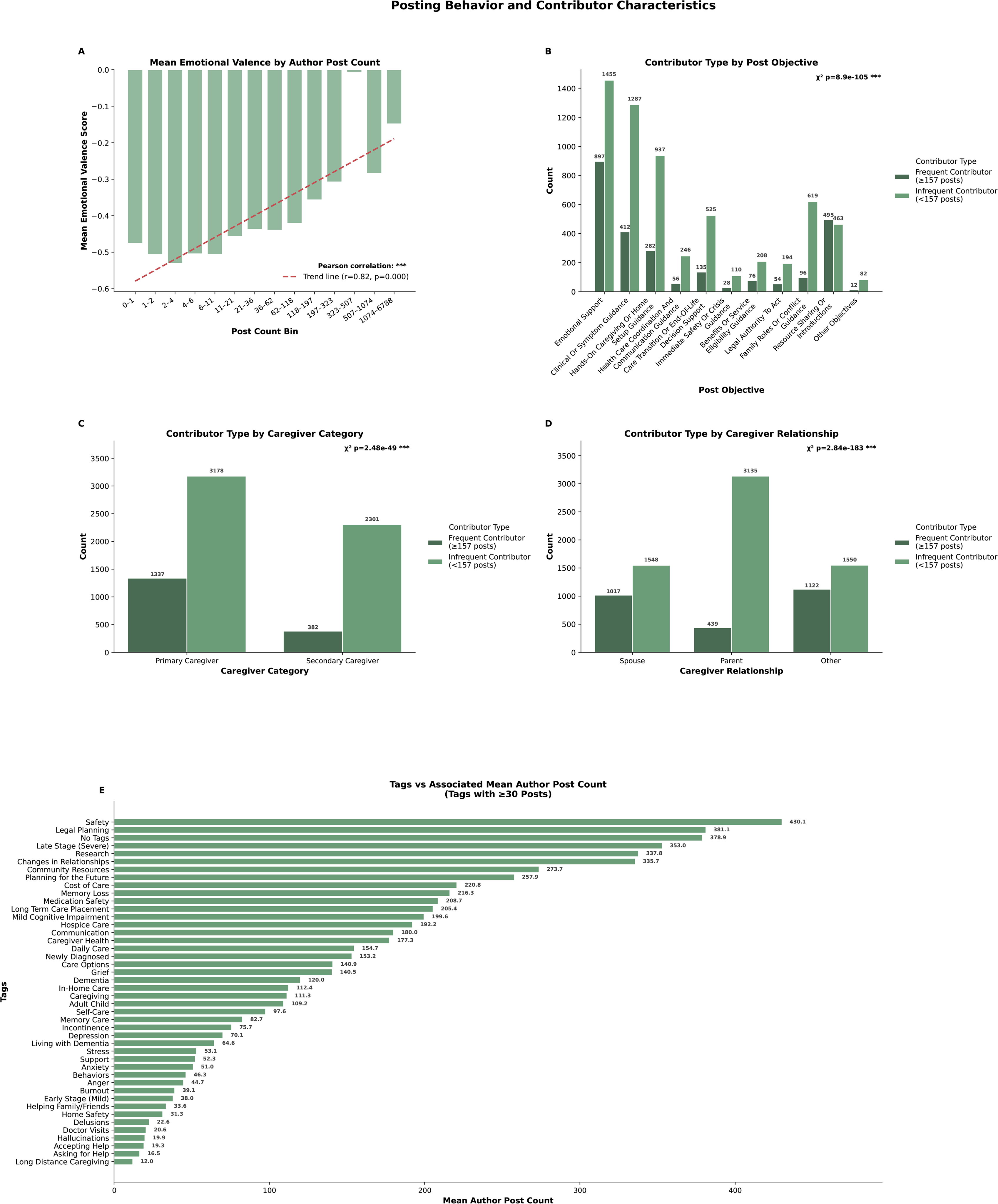
Associations between posting behavior, caregiver characteristics, and emotional expression. Mean emotional valence by author posting frequency, stratified by post-count bins, with linear trend analysis. (B) Distribution of caregiving objectives among frequent contributors (≥157 posts) and infrequent contributors (<157 posts). (C) Distribution of contributor type across caregiver status categories. (D) Distribution of contributor type across caregiver relationship categories. (E) Mean author post count associated with discussion tags appearing in at least 30 posts. Frequent contributors were defined as authors within the upper quartile of posting activity. Statistical significance for categorical comparisons was assessed using chi-square tests, and the association between posting frequency and emotional valence was evaluated using Pearson correlation. **p<0.001.

Cumulative platform posting activity was highly right-skewed (mean = 217, SD = 641), indicating that a minority of authors produced a disproportionate share of activity. Authors in the fourth quartile (75th percentile) contributed 157 or more posts and were classified as frequent contributors. Frequent contributors (≥157 posts) more often posted about resource sharing (n=495), whereas infrequent contributors more commonly posted about symptom guidance (n=1,287), hands on caregiving (n=937) and family roles and conflicts guidance (n=619) (Figure 4B).

Infrequent contributors comprised the majority of both primary (n=3178 vs 1337) and secondary caregivers (n=2301 vs 382). However, primary caregivers showed a substantially higher proportion of frequent contributors relative to secondary caregivers (n=1337 vs 382; Figure 4C). Similarly, infrequent contributors predominated across all relationship groups - parent (n=3135 vs 439), the relative proportion of frequent versus infrequent contributors varied significantly by caregiver relationship type (Figure 4D). Specifically, caregivers for spouses (n=1017) and other groups (n=1122) showed comparatively larger proportions of frequent contributors than caregivers for parents (n=439). Topics associated with the greatest posting activity (Figure 4E) included safety (mean=430), legal planning (mean=381), late stage disease (mean=353), research (mean=337), and relationship changes (mean=335).

## Discussion

Operating on a large corpus of naturally occurring posts from an online dementia caregiver community, the developed framework jointly extracted caregiver role and relationship, caregiving objective, emotional valence, and emotional themes, and linked these to community engagement. It revealed a predominantly negative emotional landscape (∼89% of posts negative; mean valence -0.46). More informatively, emotional valence differed by caregiving relationship and varied most sharply by the purpose of a post, with crisis, family conflict, and end-of-life situations the most negative and resource-sharing the only net-positive context. The central key contribution is substantive where relationship and post objective jointly account for much of the variation in caregiver emotional valence, and representing role, relationship, objective, valence, and theme together in a single structured profile makes this patterning visible in a way that a single sentiment score cannot.

A primary finding was that caregiver emotional expression was rarely reducible to a single valence label: posts classified as net positive frequently retained explicit concern alongside hope and gratitude rather than expressing uniformly positive affect, suggesting that distress and adaptive, resilience-related emotions coexist rather than alternate across the caregiving trajectory.^21,22^ This coexistence is precisely what coarse, single-label polarity classification is not positioned to capture, since it assigns each post to one category; a structured, theme-level representation instead allows valence, theme, and context to be examined jointly.

By caregiving relationship, adult-child and parent-caregivers expressed more negative valence than spouse caregivers, with adult children caring for fathers and mothers. Unlike most spouse caregivers, adult children commonly provide care while balancing employment and childrearing the “sandwich generation” and the resulting role conflict and time strain are associated with greater psychological distress.^18,19,23^ Adult children also frequently coordinate care across households and at a distance, adding logistical and communication demands, and may experience anticipatory grief as they witness a parent’s decline while negotiating shifting family roles and decision-making authority.^18,19^ Consistent with this, parent-caregivers in our data contributed disproportionately to family conflict, healthcare coordination, and legal planning discussions. Spouse caregivers, by contrast, were concentrated in emotional support and symptom management posts, patterns compatible with greater day-to-day caregiving proximity but fewer competing external obligations.

Emotional valence varied most strongly by the objective of the post. Immediate safety or crisis guidance, family roles or conflict guidance, and care transition or end-of-life decision support were the most negative contexts, similar to prior work identifying behavioral crises, safety concerns, family conflict, and care transitions as major sources of caregiver stress.^24,25^ Resource-sharing was the only objective associated with net positive expression.

Taken together, these findings indicate that expressed caregiver distress is not uniform: it varies systematically by who is caregiving and by what a post is trying to accomplish, echoing the broader literature on the psychological toll of dementia caregiving.^1,2,5,24,26,27^ It is important to be precise about what this measure represents, however: emotional valence reflects the affective tone expressed in a semi-public narrative and is shaped by the writer’s state, the purpose of the post, and self-presentation. It is one observable dimension of caregiver experience and should not be equated with clinically assessed caregiver burden or psychological distress.

The emotional theme analysis confirmed this. Concern, frustration, sadness, uncertainty, and anxiety were the dominant constructs, while guilt and distress, though less frequent, were clinically salient. The prominence of concern and uncertainty, especially within symptom-management discussions, suggests that much negative expression is anticipatory, tied to evolving disease progression and future caregiving responsibilities rather than only to present circumstances.

One of the striking observations was that emotional valence was positively associated with cumulative posting activity. More prolific contributors expressed more positive valence on average. Non-exclusive explanations and the cross-sectional design surveys cannot adjudicate among them. Sustained participation may confer benefit through accumulated social support, informational exchange, normalization of difficult experiences, and the development of coping resources such that engaged caregivers genuinely express more positive affect over time (adaptation and resilience).^21^ Alternatively, the association may reflect selection rather than change: caregivers who find the community supportive are more likely to remain and post repeatedly, whereas the acutely overwhelmed may post once and disengage, and many caregivers exit the forum at the most distressing junctures such as bereavement or when caregiving ends.

As a growing number of caregivers turn to online communities to find others navigating similar circumstances, these results point toward translational applications, while remaining a complement to, not a replacement for established assessment. A framework that derives structured caregiver phenotypes from naturally occurring text could support population-level surveillance of caregiver distress; early identification of high-risk situations, given specific objectives (crisis, family conflict, end-of-life decision making) were reliably associated with the most negative expression. It could serve as triage signals or relationship-tailored interventions, since the needs and stressors of adult-child and spouse caregivers differed systematically. Additionally, it could serve longitudinal monitoring of a community’s emotional themes and evaluation of caregiver-support programs subjected to prospective evaluation.

This study has several strengths. It draws on naturally occurring narratives rather than retrospective self-report, capturing spontaneous expression at scale. It pairs the language-model annotations with a traditional NLP comparison and assesses reproducibility through agreement between independent models, supporting the consistency of the extracted measures. And its relationship-aware design allows emotional expression to be examined jointly with role and objective rather than in isolation. Together these features offer a scalable methodology for studying caregiver experience from real-world digital interaction.

Several limitations should be acknowledged. First, the data derive from a single, English-language online platform and likely overrepresent digitally engaged, and possibly more highly educated, caregivers while underrepresenting those with limited internet access or who do not participate in online communities; participation may further favor more emotionally expressive caregivers, limiting generalizability.^28^ Second, caregiving roles and relationships were inferred from self-reported narrative rather than verified records, and relationship is partly confounded with forum membership; the most recent posts also span somewhat different calendar windows across forums. Third, expressed emotional valence is not a validated measure of caregiver burden or clinical distress, and the associations reported here are descriptive. Fourth, posts are nested within authors and a minority of authors contributed disproportionately, so post-level comparisons may overstate precision. Finally, language-model annotations remain sensitive to prompt design and model choice. Accordingly, findings should be interpreted as exploratory and complementary to survey- and interview-based research.

In conclusion, this work demonstrates that LLMs can convert unstructured caregiver narratives into structured, multidimensional phenotypes - relationship, objective, emotional valence, and emotional themes. In doing so, it reveals interpretable structure in how caregiver experience is expressed across relationships and caregiving contexts. By enabling continuous, scalable characterization of caregiver experience from naturally occurring text, this approach complements the episodic, instrument-based methods that remain the foundation of caregiving research and offers a basis for future tools to monitor caregiver experience and identify those in greatest need of support.

## Data Availability

All data produced are available online at https://alzconnected.org

## Supplemental

### Methods

#### M1: Prompts

*Prompt 1 (P1): Caregiver Status and Relationship (General Forum)*

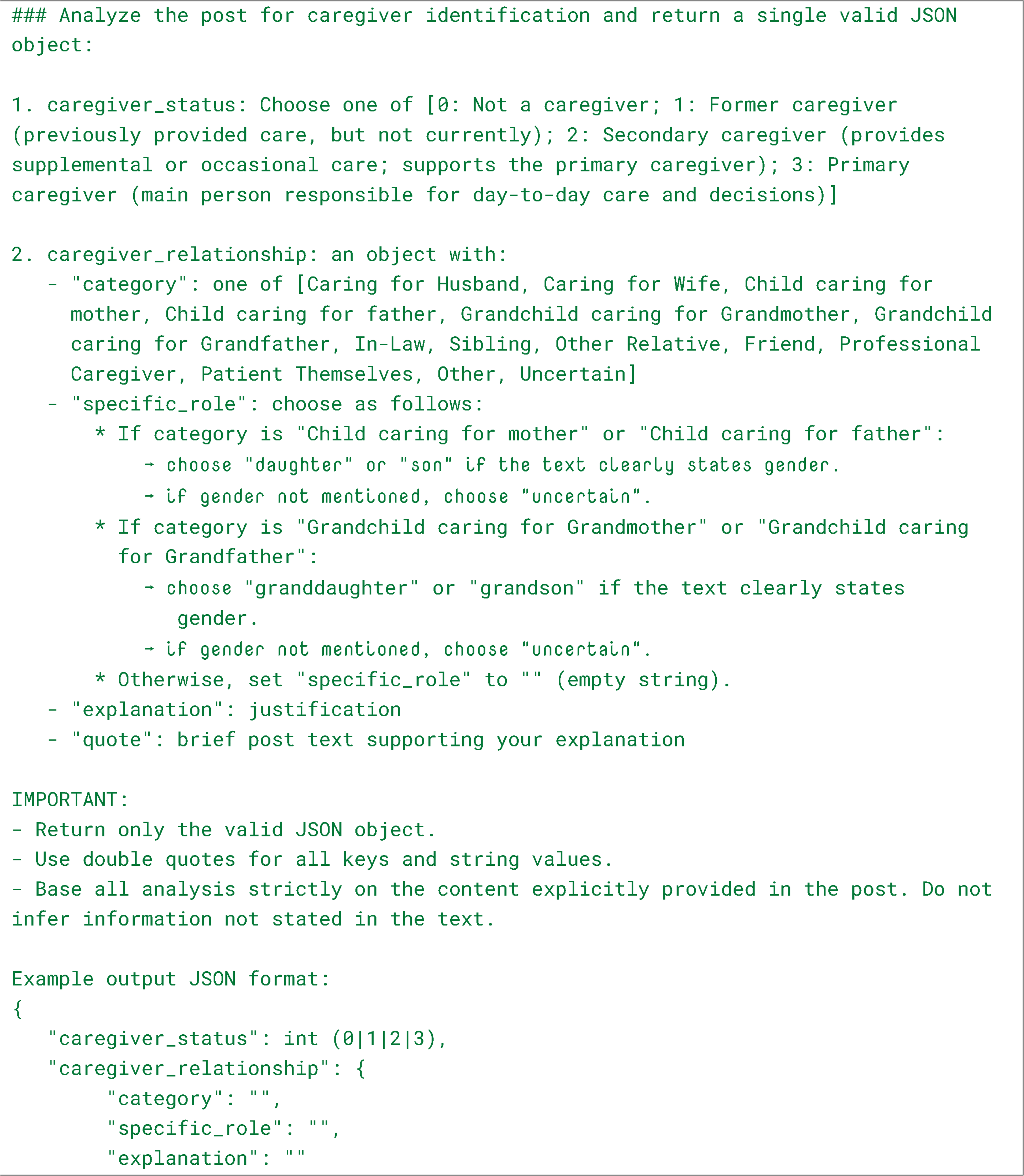
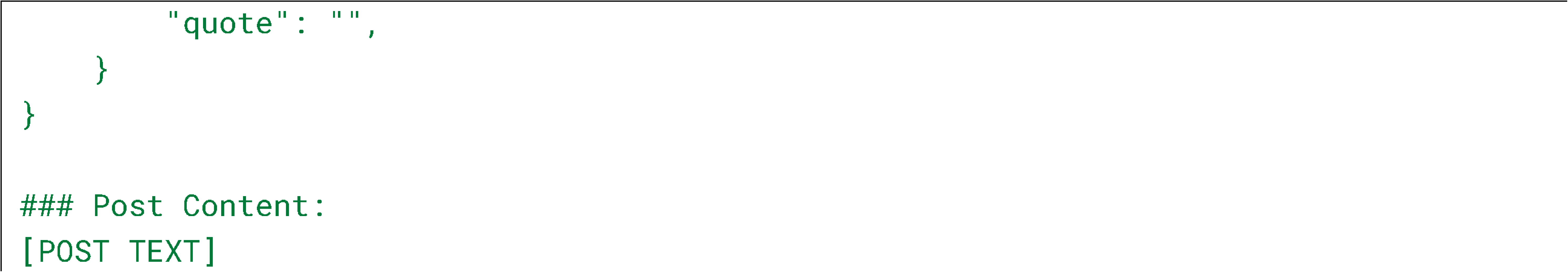

*Prompt 2 (P2): Post Objective*

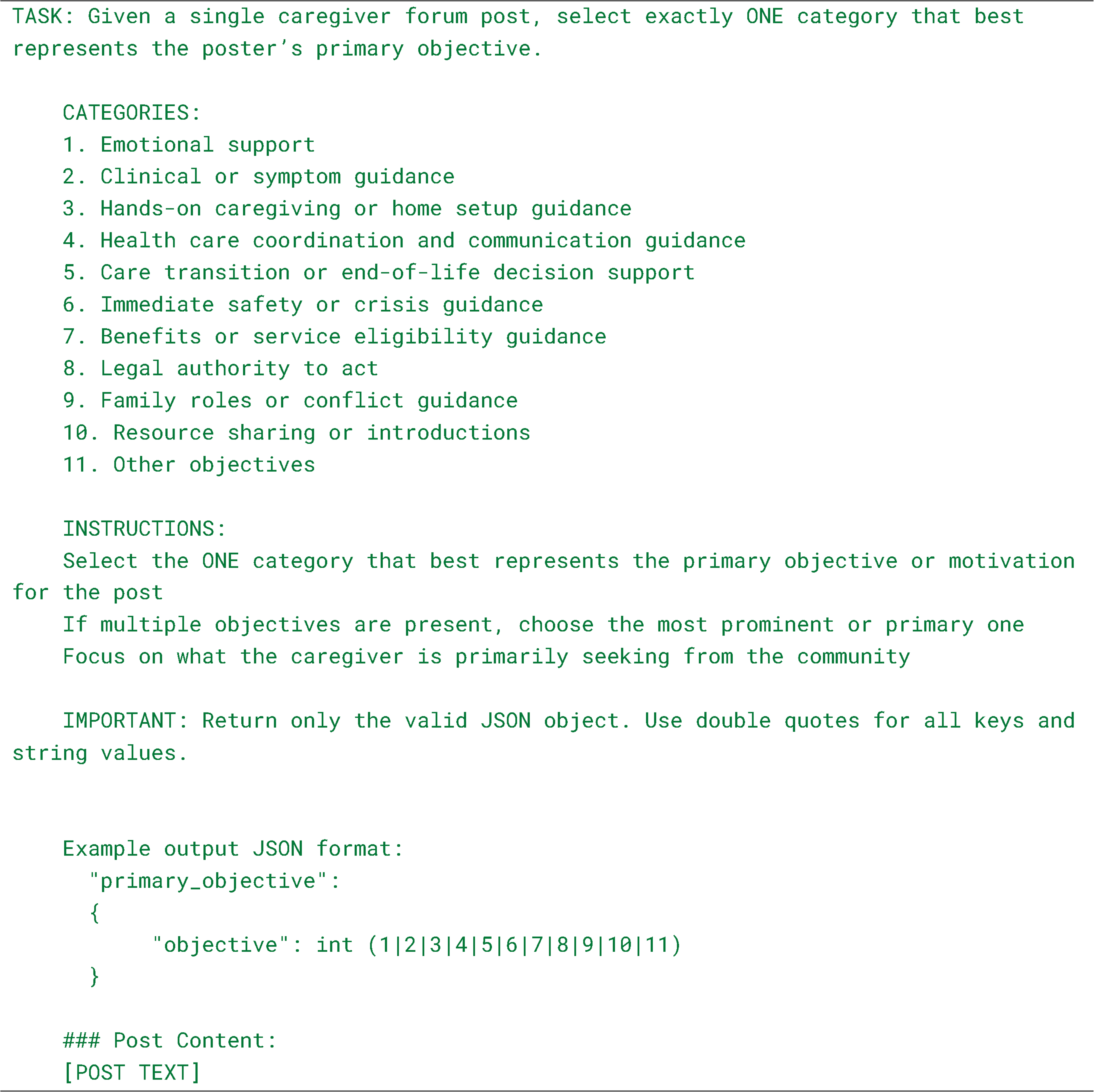

*Prompt 3 (P3): Emotional Tone*

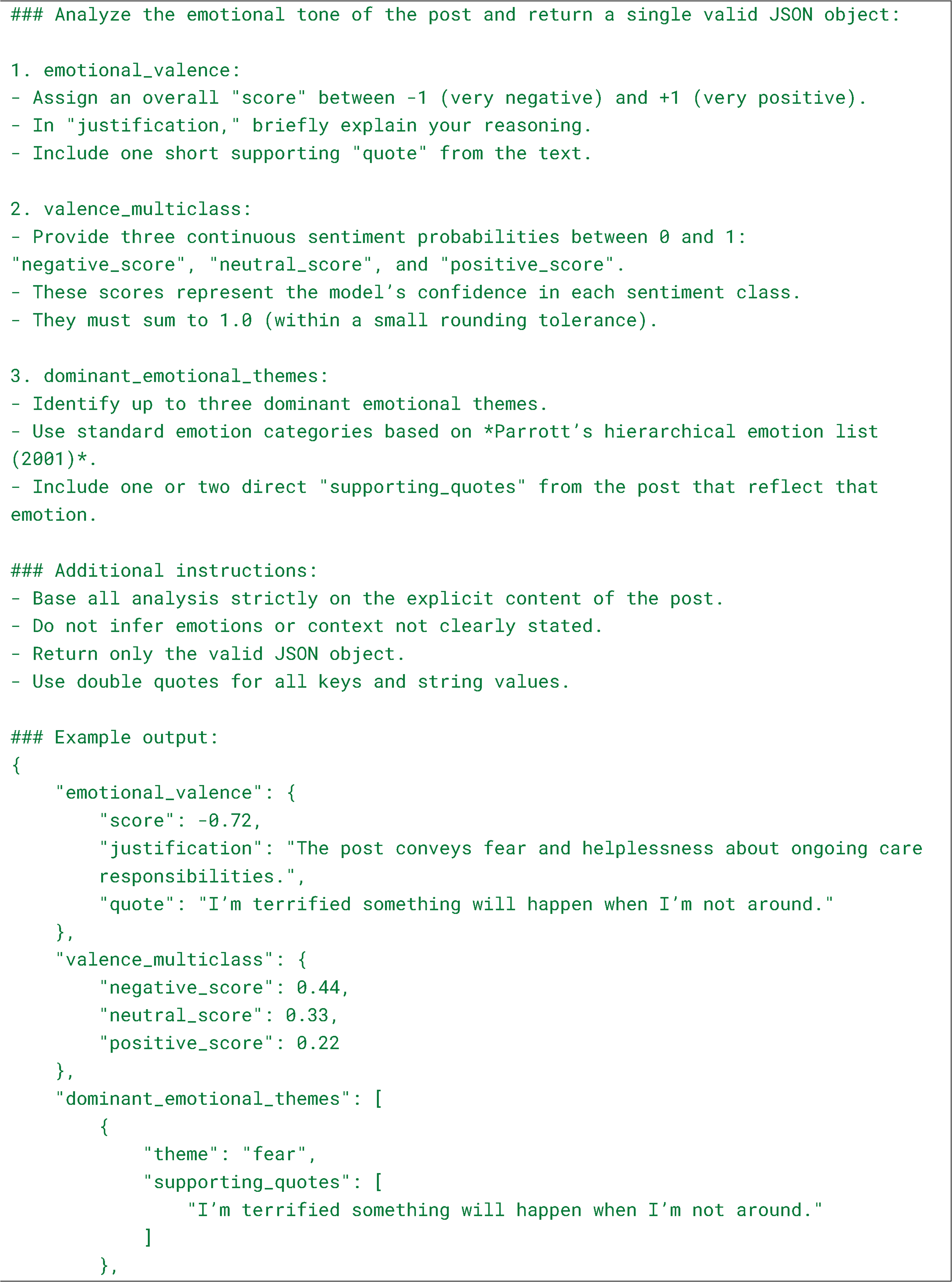
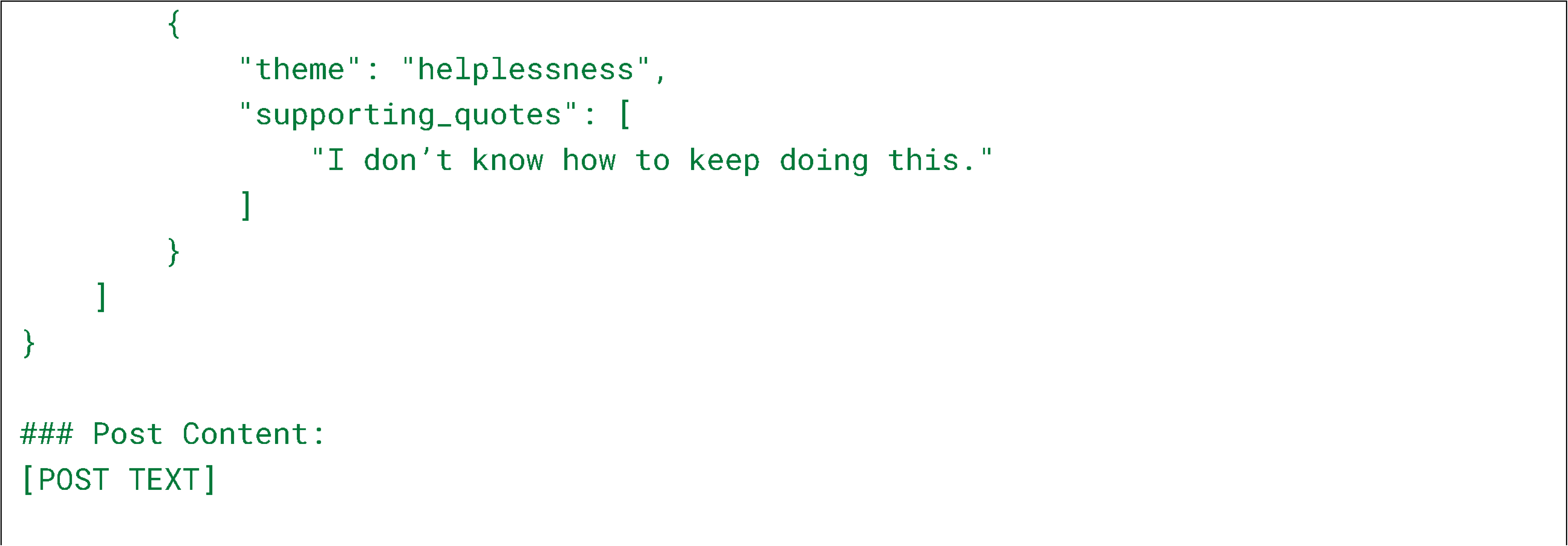

*Prompt 4 (P4): Caregiver Status and Relationship (Parent Forum)*

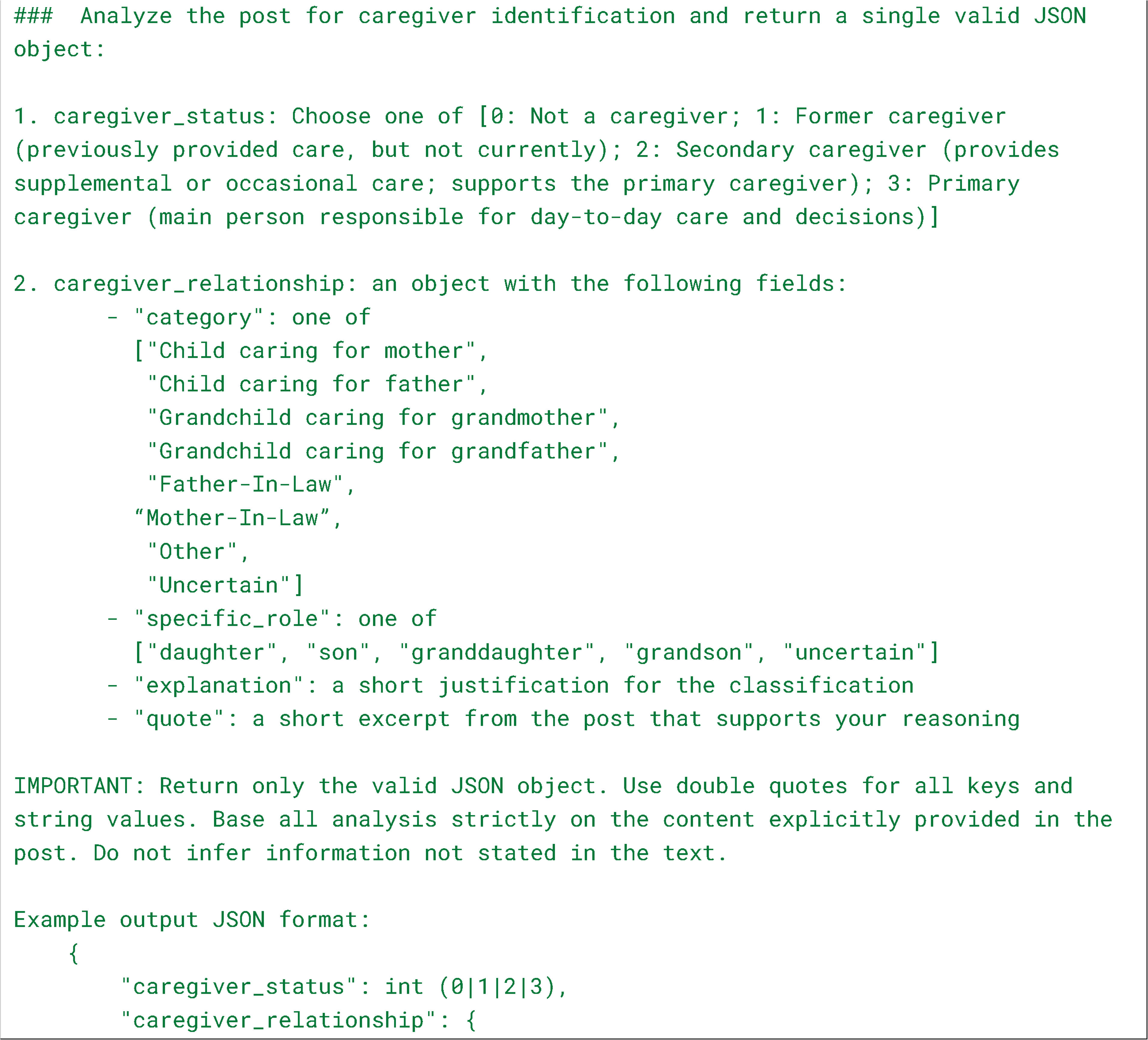
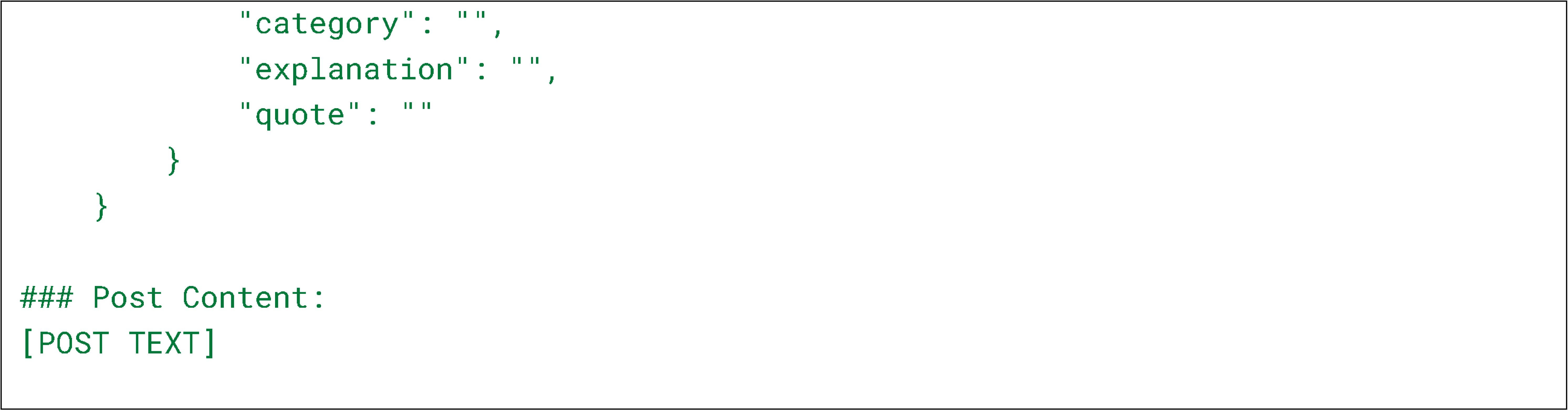

*Prompt 5 (P5): Caregiver Status and Relationship (Spouse Forum)*

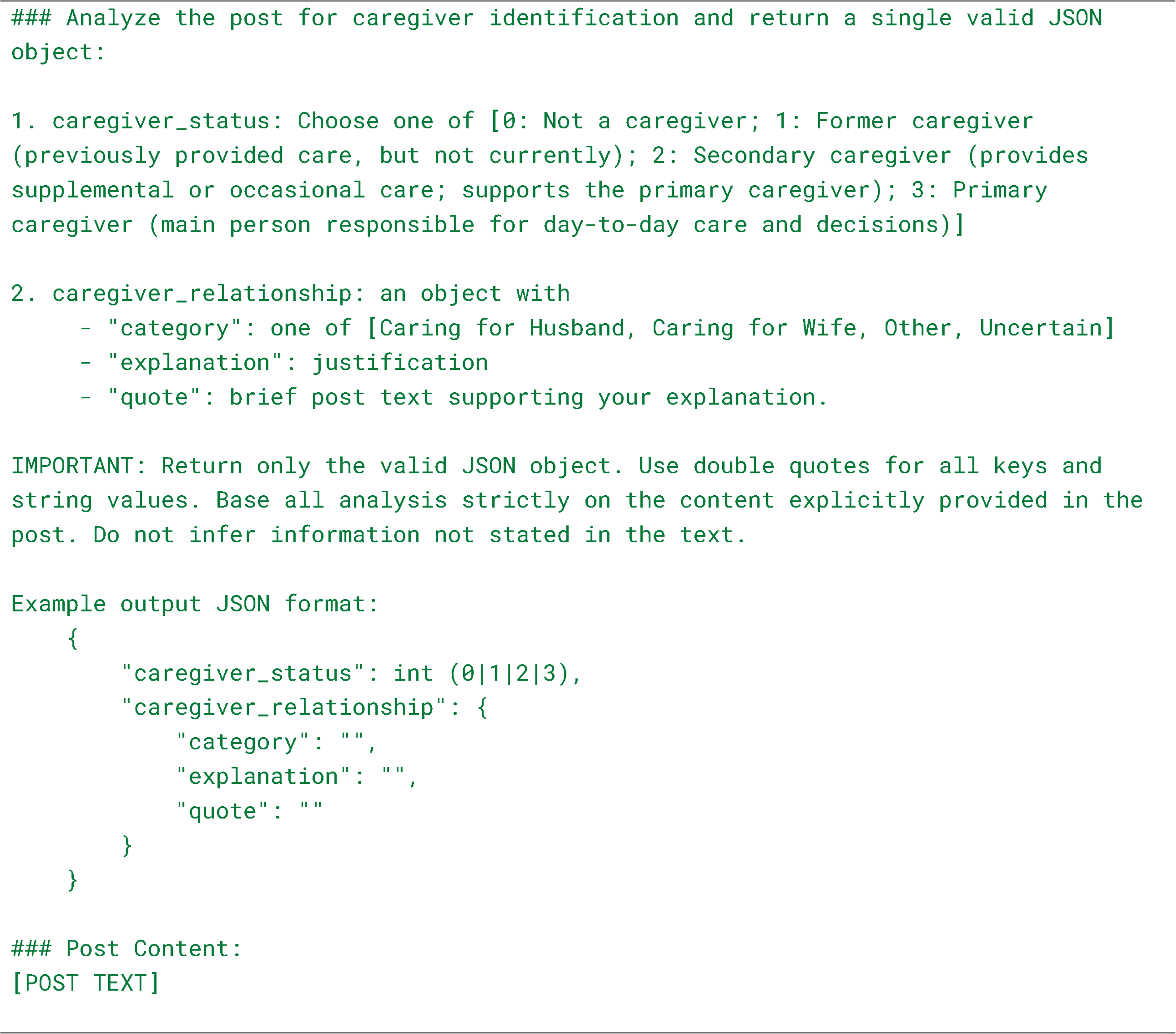

#### M2: Sentiment Analysis

##### Sentiment Analysis and Emotional Themes

Sentiment analysis was performed using a hybrid framework integrating transformer-based sentiment modeling with LLM-derived emotional annotations. Post underwent preprocessing including lowercasing, removal of hyperlinks and special characters, stop-word filtering, and lemmatization to improve textual consistency. Traditional sentiment analysis was performed using a pre-trained transformer (Supplemental Methods M2(a)) to estimate probabilities for negative, neutral, and positive emotional states, from which a weighted sentiment score was computed using class-specific values of −1, 0, and +1, respectively. These scores were compared with LLM-derived emotional valence scores, which additionally captured dominant emotional themes, supporting quotations, and explanatory statements. Histograms and distributional analyses were generated to visualize the distribution of sentiment scores and emotional valence across caregiver narratives.

##### M2(a)

Sentiment analysis was performed using the pre-trained RoBERTa-large model (j-hartmann/sentiment-roberta-large-english-3-classes; Hartmann et al., 2022). For each post, probabilities for negative, neutral, and positive sentiment were generated. A continuous sentiment score ranging from −1 to +1 was computed using weighted averaging, where negative, neutral, and positive sentiment probabilities were assigned values of −1, 0, and +1, respectively. These sentiment scores were used for distributional visualization and comparison with LLM-derived emotional valence scores.

**Figure S1.**
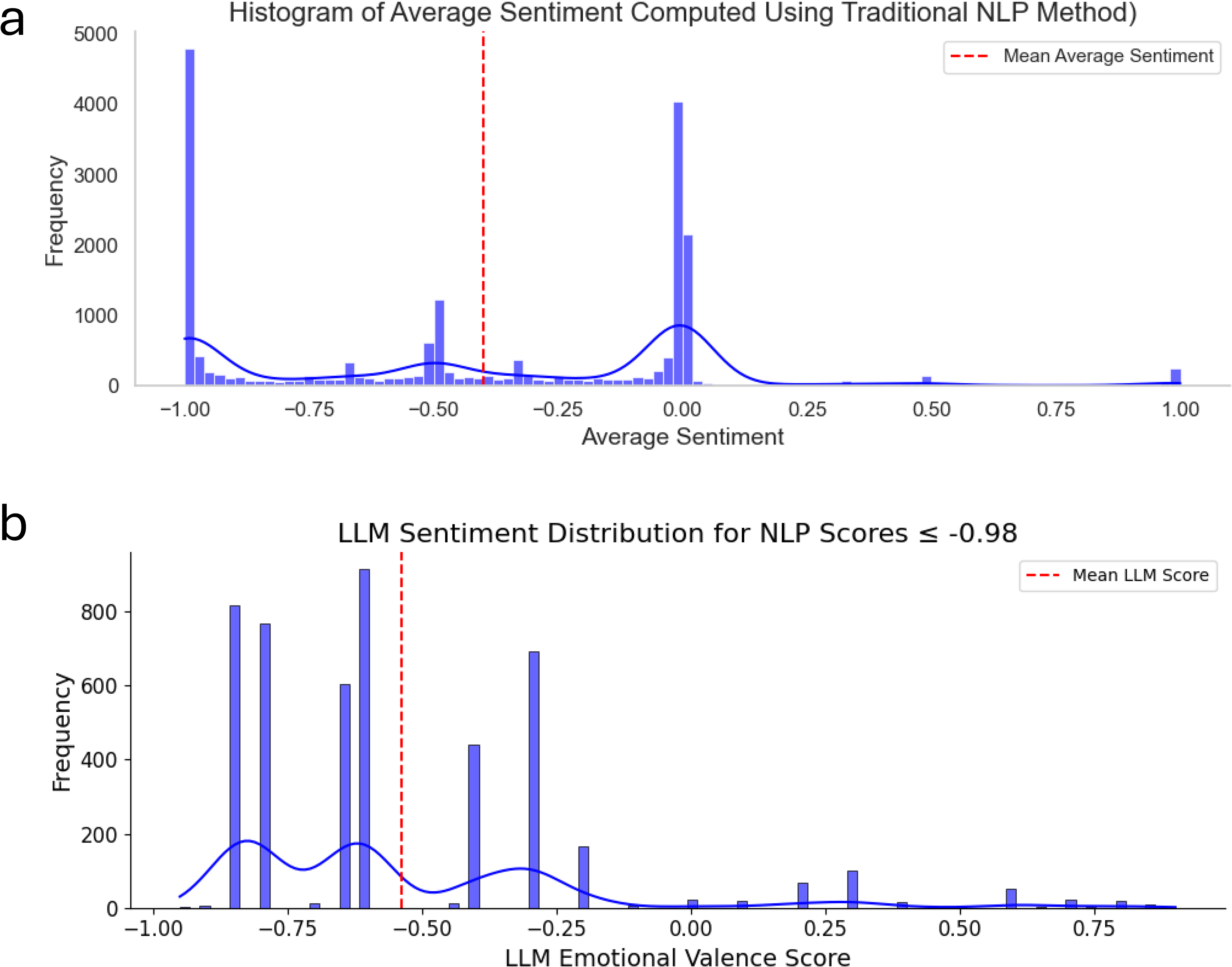
Comparison of transformer-based sentiment analysis and LLM-derived emotional valence distributions. (a) Distribution of sentiment scores generated using a pretrained RoBERTa sentiment classifier across all caregiver posts. Sentiment scores ranged from −1 (most negative) to +1 (most positive), with the red dashed line indicating the mean sentiment score. (b) Distribution of LLM-derived emotional valence scores among posts classified as extremely negative by the transformer model (sentiment score ≤ −0.98). The red dashed line indicates the mean LLM emotional valence score.

